# Adaptability of the Pulsta valve to the diverse main pulmonary artery shape of native right ventricular outflow tract disease

**DOI:** 10.1101/2023.11.20.23298796

**Authors:** Woo Young Park, Gi Beom Kim, Sang Yun Lee, Ah Young Kim, Jae Young Choi, So Ick Chang, Seong Ho Kim, Seul Gi Cha, Jou-Kou Wang, Ming-Tai Lin, Chun-An Chen

## Abstract

**Background:** Pulsta valve is increasingly used for percutaneous pulmonary valve implantation (PPVI) in patients with large native right ventricular outflow tract (RVOT). The aim of this study is to reveal Pulsta valve implantation outcomes within the native RVOT and to evaluate the adaptability of Pulsta valve to diverse native main pulmonary artery (PA).

**Methods:** A multicenter retrospective study encompassing 183 patients with moderate to severe pulmonary regurgitation (PR) in the native RVOT who underwent PPVI with Pulsta valves® between February 2016 and August 2023 at the five Korean and Taiwanese tertiary referral centers.

**Results:** Successful implantation of the Pulsta valves was achieved in 180 out of 183 patients (98.4 %) with an average age of 26.6 ± 11.0 years. Mean follow-up duration was 29 months. Baseline assessments revealed enlarged right ventricle (RV) volume (mean indexed RV end-diastolic volume: 163.1(Interquartile range, IQR, 152.2-179.9) mL/m^2^), which significantly decreased to 123.0(IQR: 106.9-137.2) mL/m^2^ after one year. In this study, the main PA types were classified as follows: pyramidal (3.8%), straight (38.3%), reverse pyramidal (13.7%), convex (26.2%), and concave (18.0%) shapes. Pulsta valve placement was adapted, with distal main PA for pyramidal shapes and proximal or mid-PA for reverse pyramidal shapes. The remaining patients underwent Pulsta valve implantation in the proximal or mid part of the main PA, depending on the anatomical features and size of the main PA. Two patients experienced Pulsta valve embolization to RV, necessitating surgical removal, and one patient encountered valve migration to the distal main PA, necessitating surgical fixation.

**Conclusions:** Customized valve insertion sites are pivotal in self-expandable PPVI considering diverse native RVOT shape. Rather soft and compact structure of Pulsta valve has characteristics to be adaptable to diverse native RVOT geometries.

**What is Known?:** The Pulsta valve is a self-expandable knitted nitinol-wire stent mounted with a treated tri-leaflet α-Gal-free porcine pericardial valve for percutaneous pulmonary valve implantation in patients with native right ventricular outflow tract lesions.

**What the study adds?:** To this date, this study is the largest study among the previous reports that examined the outcomes of the Pulsta valve. The Pulsta valve consistently demonstrated favorable clinical and hemodynamic outcomes over a 2.4-year period. Pulsta valve showed adaptability in the diverse native right ventricular outflow tract geometries from the compact shape of Pulsta valve.

## Introduction

Percutaneous pulmonary valve implantation (PPVI) has emerged as a minimally invasive, transcatheter-based approach aimed at prolonging the lifespan of conduits/homografts by addressing stenosis and regurgitation without the need for open-heart surgery.^1^ The pioneering work of Bonhoeffer et al. in 2000 marked the first successful human PPVI, subsequently demonstrating its safety and efficacy in selected patients with right ventricular outflow tract (RVOT) dysfunction.^2–4^

Balloon-expandable PPVI using Melody and Edwards SAPIEN valves has been most common method so far. However, balloon-expandable PPVI has certain limitations when dealing with native RVOT lesions characterized by varying geometries and significant pulmonary regurgitation.^5^ Consequently, a larger self-expandable valve has been proposed as the next generation of percutaneous valves for these native RVOT lesions.^6–10^ One such valve is the Pulsta valve (TaeWoong Medical Co, Gyeonggi-do, South Korea). The Pulsta valve is designed as a self-expandable valve with symmetrical flaring on both ends, enabling it to accommodate the larger dimensions of the native RVOT. Furthermore, it utilizes a relatively low-profile delivery catheter.^10^ The introduction of the Pulsta valve has expanded the indications for PPVI to include patients with large native RVOTs.

However, it is important to note that the native RVOT exhibits diverse anatomical structures in each patient. Consequently, during the performance of PPVI, the choice of insertion site must be individualized according to the patient’s specific RVOT morphology. The objective of this study is to present the outcomes of the Pulsta valve implantation from multi-center data and propose an insertion site based on the anatomy of the main pulmonary artery (PA) using Pulsta valve.

## Methods

### Patients

We conducted a multi-center, retrospective study of the 183 patients with moderate to severe pulmonary regurgitation (PR) within the native RVOT. These patients underwent PPVI with a Pulsta valve® at the four pediatric cardiac centers in South Korea and an additional pediatric cardiac center in Taiwan at any time between January 2016 and August 2023. The patients who underwent valve-in-valve procedures were excluded.

### Device Design

The Pulsta valve is a self-expandable tri-leaflet valve using treated porcine pericardium which is basically open state. (Figure 1, Supplement video 1) The Pulsta valve exhibits a cylindrical shape with uncovered proximal and distal ends, enabling precise positioning without compromising blood flow. The valve diameter ranges from 18 to 32 mm with 2 mm increments. Both ends of the valve are flared 4 mm wider than the outer diameter. The total length of the valve is 28∼38 mm according to outer diameter. The Pulsta valve is using a relatively low-profile delivery catheter. The diameter of the outer sheath at the head portion is 18 French up to 28 mm valve and 20 French for 30 and 32 mm valve. In addition, there is 3 hook blocks at the head portion for stable deployment.

**Figure 1.**
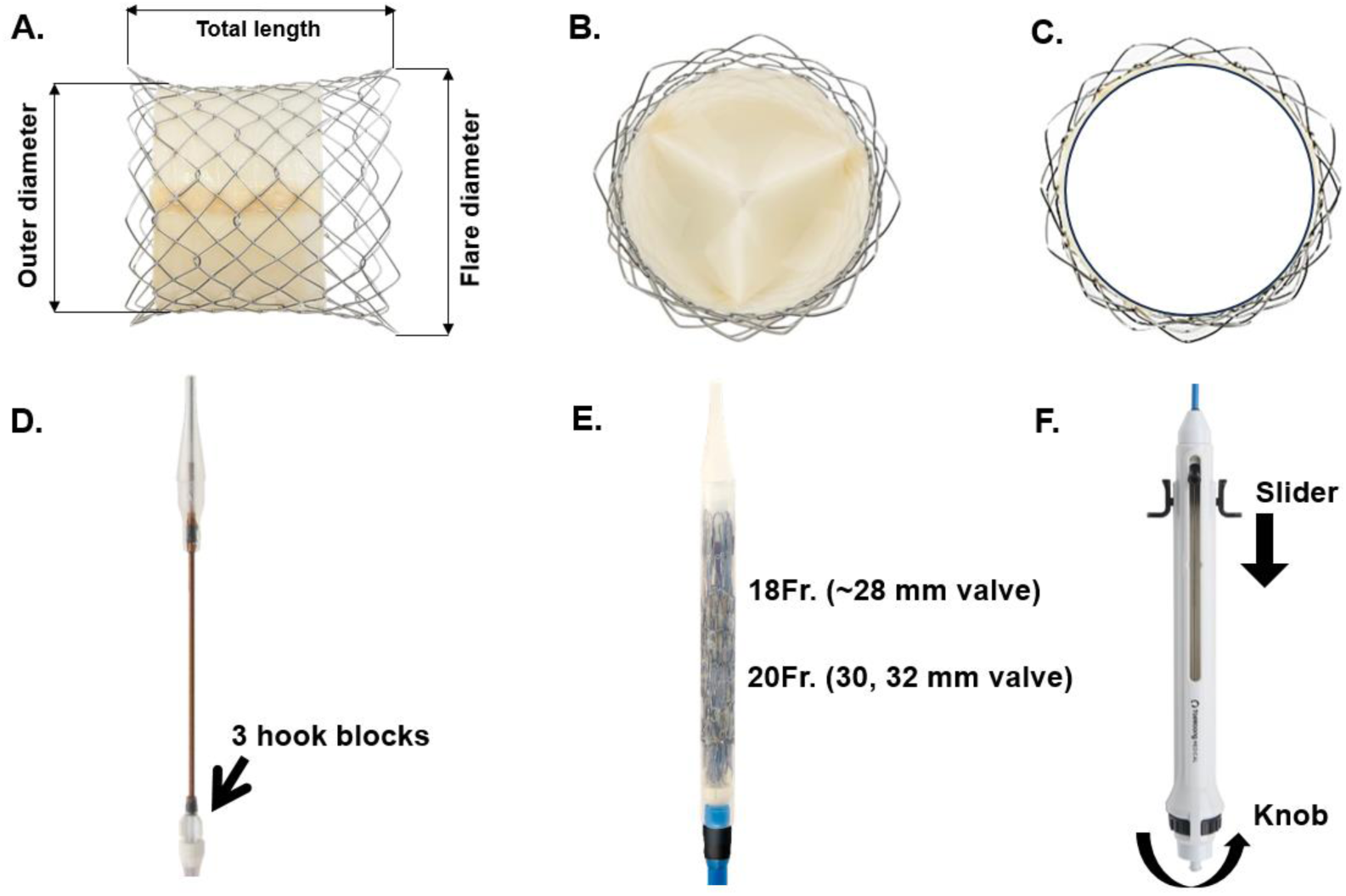
Pulsta valve profile and delivery cable catheter system (A) The Pulsta valve exhibits a cylindrical shape with uncovered proximal and distal ends. The valve diameter ranges from 18 to 32 mm with 2 mm increments. Both ends of the valve are flared 4 mm wider than the outer diameter. The total length of the valve is 28–38 mm according to outer diameter. (B, C)This tri-leaflet valve is made using treated porcine pericardium and is essentially an open-state valve. (D) A hook block is incorporated at the head portion to prevent valve jumping during full deployment. (E)The diameter of the outer sheath at the head portion is 18 French up to 28 mm valve and 20 French for 30 and 32 mm valve. (F) By rolling the knob clockwise, the outer sheath is pulled back and half of the valve can be expanded, and the valve is completely expanded by pulling the slider.

### RVOT type definition

The RVOT of the patients was classified according to the classification presented by S.

Schievano in 2007.^11^ We defined each RVOT type as below for this study.

Type 1. Pyramidal: proximal PA annulus diameter ≥ 1.3 X distal bifurcation site diameter

Type 2. Straight: parallel walls of the outflow tract and widest area diameter ≤ 1.3 X proximal PA annulus or distal bifurcation site diameter

Type 3. Reverse pyramidal: distal bifurcation site diameter ≥ 1.3 X proximal PA annulus diameter

Type 4. Convex: widest area diameter ≥ 1.3 X proximal PA annulus and distal bifurcation site diameter

Type 5. Concave: proximal PA annulus and distal bifurcation site diameter ≥ 1.3 X narrowest mid PA diameter

### Pulmonary artery size measurement

In clinical practice, the target valve implantation area for PPVI is typically the main PA itself. The size measurement of the main PA is usually determined by incorporating all available imaging modalities such as transthoracic echocardiography, cardiac computed tomography, and cardiac magnetic resonance imaging (MRI) prior to the procedure. Additionally, cine angiography and balloon sizing assessment for main PA diameter and length measurement are performed during the procedure.

To obtain measurements of the target main PA, various parameters are considered during its most dilated phase. These include the proximal PA annulus, the mid or narrowest section of the main PA, the distal bifurcation site, and the length of the main PA. Measurements are obtained from multiple imaging modalities, including transthoracic echocardiography performed at the parasternal short or long axis view primarily during the mid- to end-systolic phase (the phase when the main PA is most dilated), as well as angiography performed at the right or left anterior oblique and true lateral view.

The main PA length is typically measured from the level of the proximal annulus of the main PA to the distal PA bifurcation site. By utilizing these measurements and imaging techniques, we accurately assess the dimensions of the main PA, aiding in the planning and execution of PPVI procedures.

### Valve size selection

For valve sizing in PPVI, we selected a valve size that was 2∼4 mm larger than the narrowest area of the main PA and 1∼2 mm larger than the overall size of the main PA during its maximal dilated phase observed during main PA angiography and sizing balloon measurement of main PA. During these procedures, we aimed to avoid excessive dilation of the sizing balloon to minimize any discrepancies in the measurement of the narrowest diameter of the main PA obtained from various imaging modalities.

In cases where determining the optimal valve size was challenging, we employed balloon interrogation technique using static non-compliant balloon with discrete maximal diameter such as Tyshak or BiB catheter. We could confidence to decide 1∼2 mm larger diameter valve if there is no leakage by occluding main PA landing area by static non-compliant balloon during RV angiography.

### Data collection and follow up

In this study, we documented patients’ clinical findings, echocardiographic findings, and cardiac MRI data at baseline and 6∼12 months after PPVI. Throughout the follow-up period after PPVI, we carefully checked any adverse events such as infective endocarditis, stent fracture, etc. This study was approved by the Institutional Review Board of the Seoul National University Hospital (IRB number: 2209-130-1362).

### Statistical Analysis

SPSS version 23.0 (IBM, Armonk, NY) and GraphPad Prism software were used for data analysis. For the descriptive analysis, continuous variables are described as mean ± SD or median with interquartile range, while categorical variables are presented as frequencies and percentage. The Wilcoxon signed-rank test was used to compare variables between baseline and after PPVI. Statistical significance was defined as p < 0.05.

## Results

### Study Population and characteristics

183 patients with moderate to severe PR and enlarged RV after total correction of tetralogy of Fallot, congenital pulmonary stenosis, pulmonary atresia and Fallot type double outlet right ventricle are enrolled for this study. Patient’s baseline characteristics before procedure are described in Table 1. Mean age was 26.6 ± 11.0 years (range: 10∼69 years) and mean body weight was 60.5 ± 15.3 kg (range: 28.8∼110.0). Among them, 102 patients were male (55.7%). 128 patients (80.0%) underwent transannular RVOT patch angioplasty at the time of initial total correction. Mean main PA diameter at the narrowest area was 22.9±3.4 mm (range: 11.0∼33.2) and mean main PA diameter at the widest area was 31.2±4.3 mm (range: 19.1∼51.0).

**Table 1.** Patients’ baseline characteristics before procedure.

|  |  |
| --- | --- |
| Age, years | 26.6±11.0 (10~69) |
| Gender(M/F) | 102/81 |
| Body weight, kg | 60.5±15.3 (28.8~110.0) |
| BSA | 1.6±0.2 (1.1~2.4) |
| CHDs |  |
| TOF, N (%) | 154/183 (84.2%) |
| PA, IVS, N (%) | 9/183 (4.9%) |
| PA, VSD, N (%) | 5/183 (2.7%) |
| DORV, N (%) | 6/183 (3.3%) |
| Congenital PS, N (%) | 7/183 (3.8%) |
| Others | 2/183 (1.1%) |
| Transannular RVOT widening, N (%)<sup>*</sup> | 128/160 (80.0%) |
| RVOT aneurysm, N (%) | 101/183 (55.2%) |
| LPA stenosis, N (%) | 39/183 (21.3%) |
| LPA stent, N (%) | 13/183 (7.1%) |
| RPA stenosis, N (%) | 3/183 (1.6%) |
| RPA stent, N (%) | 1/183 (0.6) |
| Main PA proximal diameter, mm | 25.4±3.9 (14.7~41.0) |
| Main PA distal diameter, mm | 27.9±5.1 (16.8~51.0) |
| Main PA length diameter, mm | 37.5±5.0 (22.0~51.9) |
| Main PA widest diameter, mm | 31.2±4.3 (19.1~51.0) |
| Main PA narrowest diameter, mm | 22.9±3.4 (11.0~33.2) |

Note: Values are number or mean ± SD (range)
Abbreviations: BSA, body surface area; CHDs, congenital heart disease; TOF, tetralogy of Fallot; PA, pulmonary atresia; VSD, ventricular septal defect; IVS, intact ventricular septum; DORV, double outlet right ventricle; PS, pulmonary stenosis; RVOT, right ventricular outflow tract; LPA, left pulmonary artery; RPA, right pulmonary artery; PA, pulmonary artery
\* 23 missing data

### Main pulmonary artery morphology and insertion site

All patients were categorized into five distinct types based on their main PA morphology. The procedural characteristics for each type are described in Table 2. Specifically, there were 7 patients (3.8%, 7/183 patients) classified as Type 1, 70 patients (38.3%, 70/183 patients) as Type 2, 25 patients (13.7%, 25/183 patients) as Type 3, 48 patients (26.2%, 48/183 patients) as Type 4, and 33 patients (18.0%, 33/183 patients) as Type 5. All patients with Type 1 underwent transannular RVOT patch angioplasty at the time of initial total correction and all of them had RVOT aneurysm. In patients with a Type 1 and Type 3 main PA shape, approximately half of the patients underwent implantation of a 32mm diameter Pulsta valve. Additionally, in all cases, a 38mm length Pulsta valve was used for implantation. This decision to use slightly larger Pulsta valves in Type 1 and Type 3 patients was made due to the higher risk of instability compared to other types.

**Table 2.**
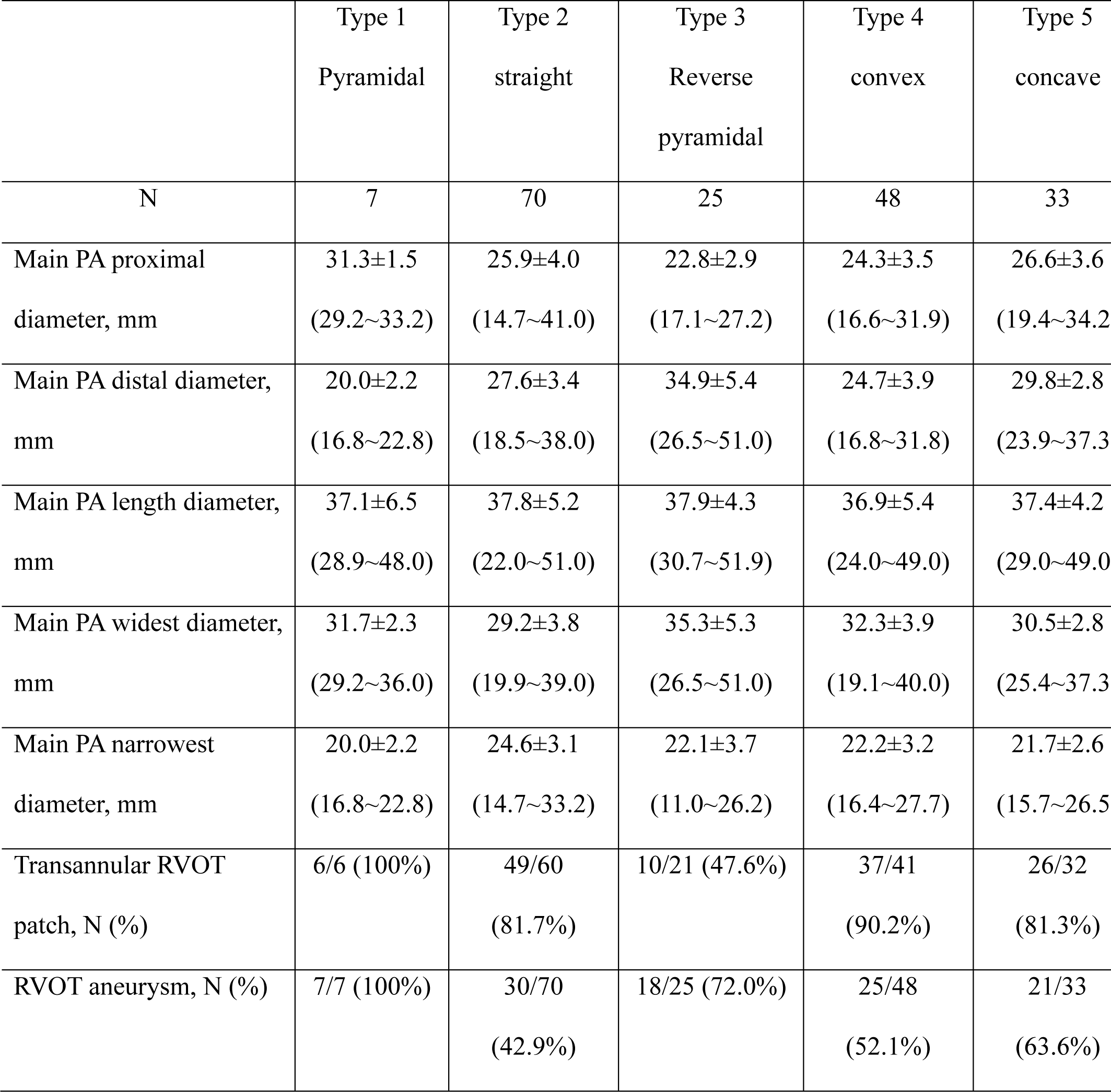
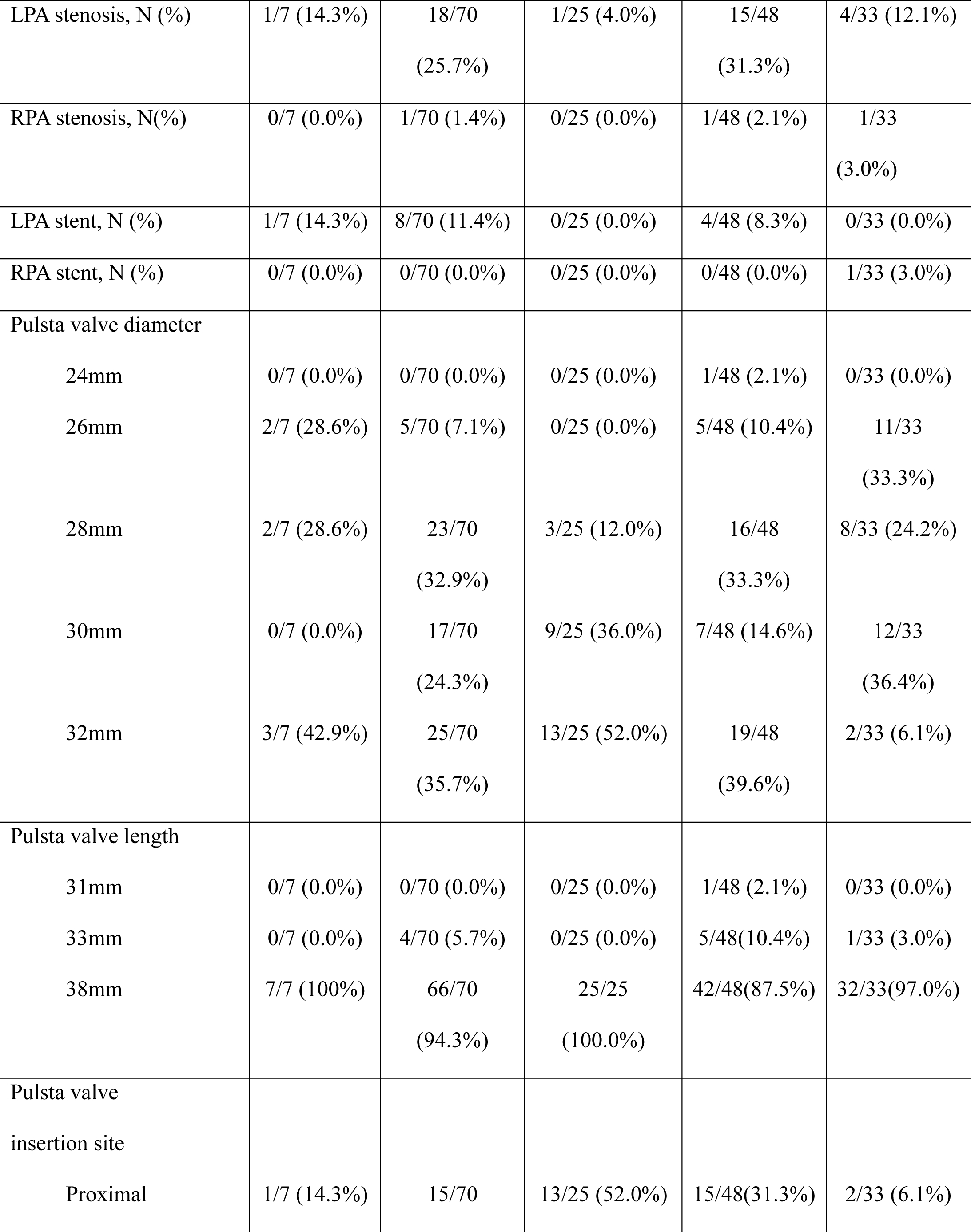
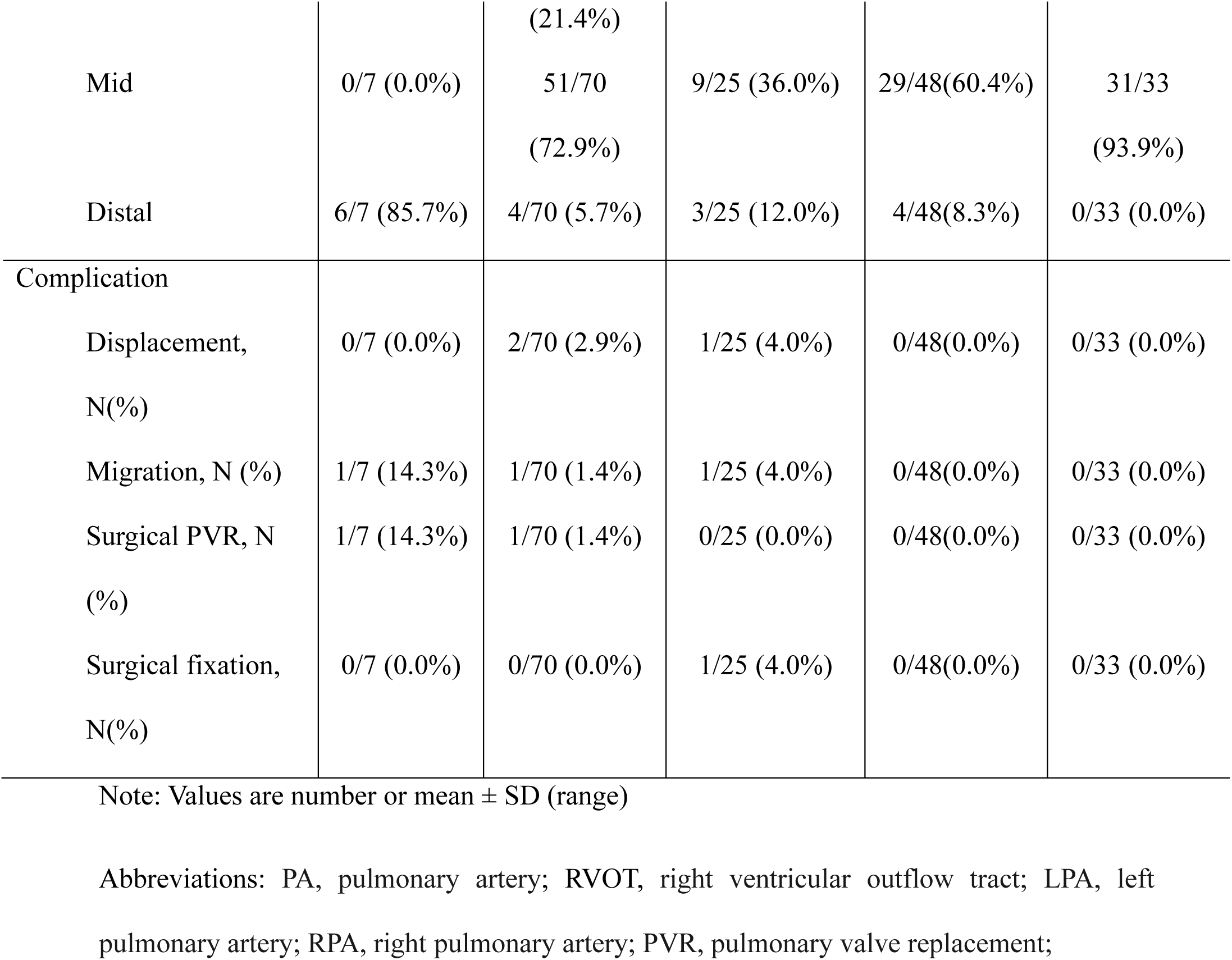
Main pulmonary artery shape and procedural success.

The insertion site of the Pulsta valve, categorized by the main PA morphology, is summarized in Figure 2. In the case of all patients except one with Type 1 main PA, PPVI was performed at the distal part of the main PA. However, in a single case, a Pulsta valve was implanted at the proximal main PA, but it subsequently embolized during the transfer to the intensive care unit post-procedure. This patient underwent surgical removal of the embolized Pulsta valve followed by surgical prosthetic pulmonary valve replacement.

**Figure 2.**
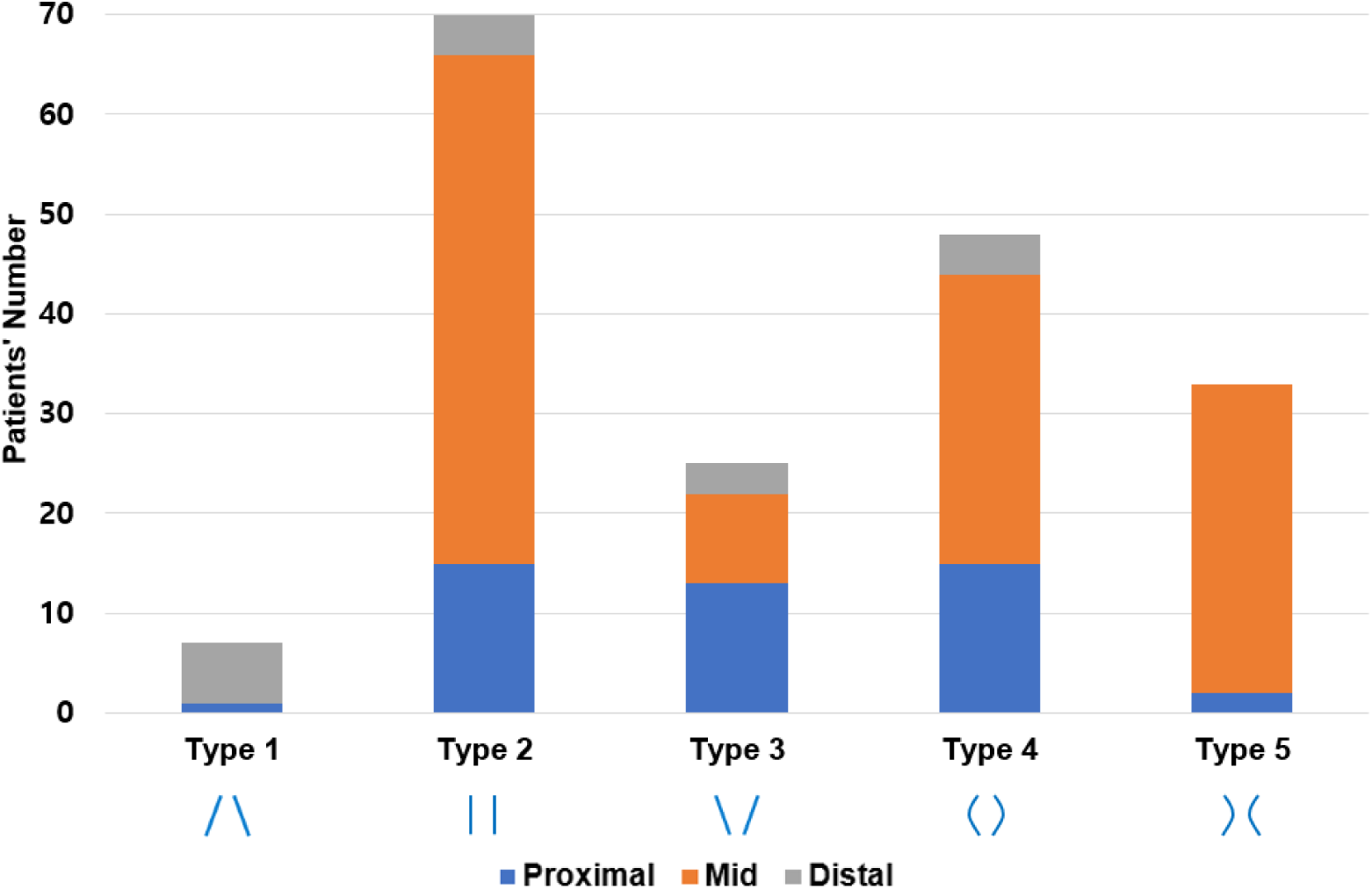
Pulsta valve insertion site status according to main pulmonary artery shape. For type 1 main pulmonary artery with pyramidal shape, Pulsta valve implantation was targeted at the distal part of the main pulmonary artery. For type 3 main pulmonary artery with reverse pyramidal shape, most Pulsta valve was implanted at the proximal narrowest area and followed at the mid part of main pulmonary artery.

Most patients with a Type 2 (72.9%) received PPVI at the mid part of the main PA. In one case of Type 2 shape, the Pulsta valve was initially implanted at the middle part of the main PA but subsequently embolized to RV due to under-sizing of valve diameter. This patient underwent surgical removal of the embolized Pulsta valve followed by surgical prosthetic pulmonary valve replacement. In another case within the Type 2 main PA shape, the Pulsta valve was initially implanted at the middle part of the main PA but subsequently displaced to the proximal part of the main PA. In response to this displacement, the patient underwent a second Pulsta valve implantation on the same day of the procedure.

The majority of type 3 patients underwent PPVI either at the proximal segment of the main PA (52.0%) or the mid part of the main PA (36.0%). In one Type 3 case, the Pulsta valve was initially implanted at the mid part of the main PA but migrated to the distal part of the main PA due to the patient’s sudden movement during the Pulsta valve’s full deployment. This patient underwent surgical fixation of the embolized Pulsta valve.

Most patients in type 4 (60.4%) and type 5 (93.9%) underwent PPVI at the mid part of the main PA.

### Follow up

We conducted a scheduled follow-up assessment for all patients over a median follow-up duration of 29 months, with a range from 0.3 to 89 months without any follow-up loss. Hemodynamic and clinical changes after Pulsta valve implantation are described in Figure 3 and Table 3. Values are reported as median and interquartile range because the normality of the distribution was rejected. Under the Wilcoxon signed-rank test, the mean pulmonary systolic pressure gradient, right ventricular end-diastolic volume index, right ventricular end-systolic volume index, right ventricular ejection fraction, and PR fraction showed statistically significant decreases after PPVI. New York heart association functional class of patients improved to class I or II in all patients.

**Figure 3.**
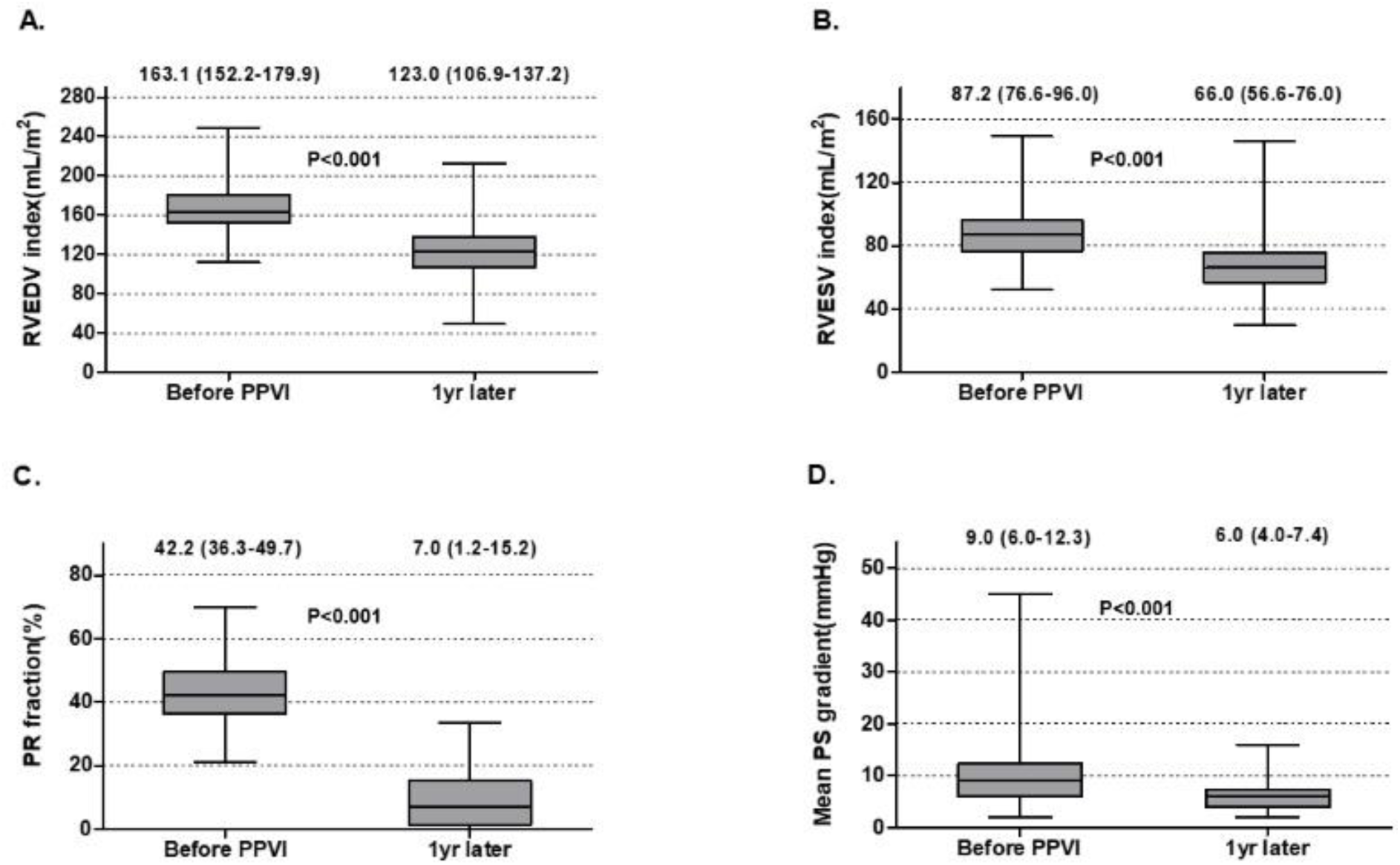
Hemodynamic changes following Pulsta valve implantation. The RV end-diastolic volume index (A), RV end-systolic volume index (B), and PR fraction (C) demonstrated statistically significant reductions after Pulsta valve implantation on Cardiac MRI. The mean PS pressure gradient (D) also demonstrated statistically significant reductions after Pulsta valve implantation on echocardiography. Values are presented as median (interquartile range). The box plot uses the median and the lower and upper quartiles (defined as the 25^th^ and 75^th^ percentiles) RV; right ventricle, RVEDV; right ventricle end-diastolic volume, RVESV; right ventricle end-systolic volume, PR; pulmonary regurgitation, MRI; magnetic resonance imaging; PS, pulmonary stenosis;

**Table 3.**
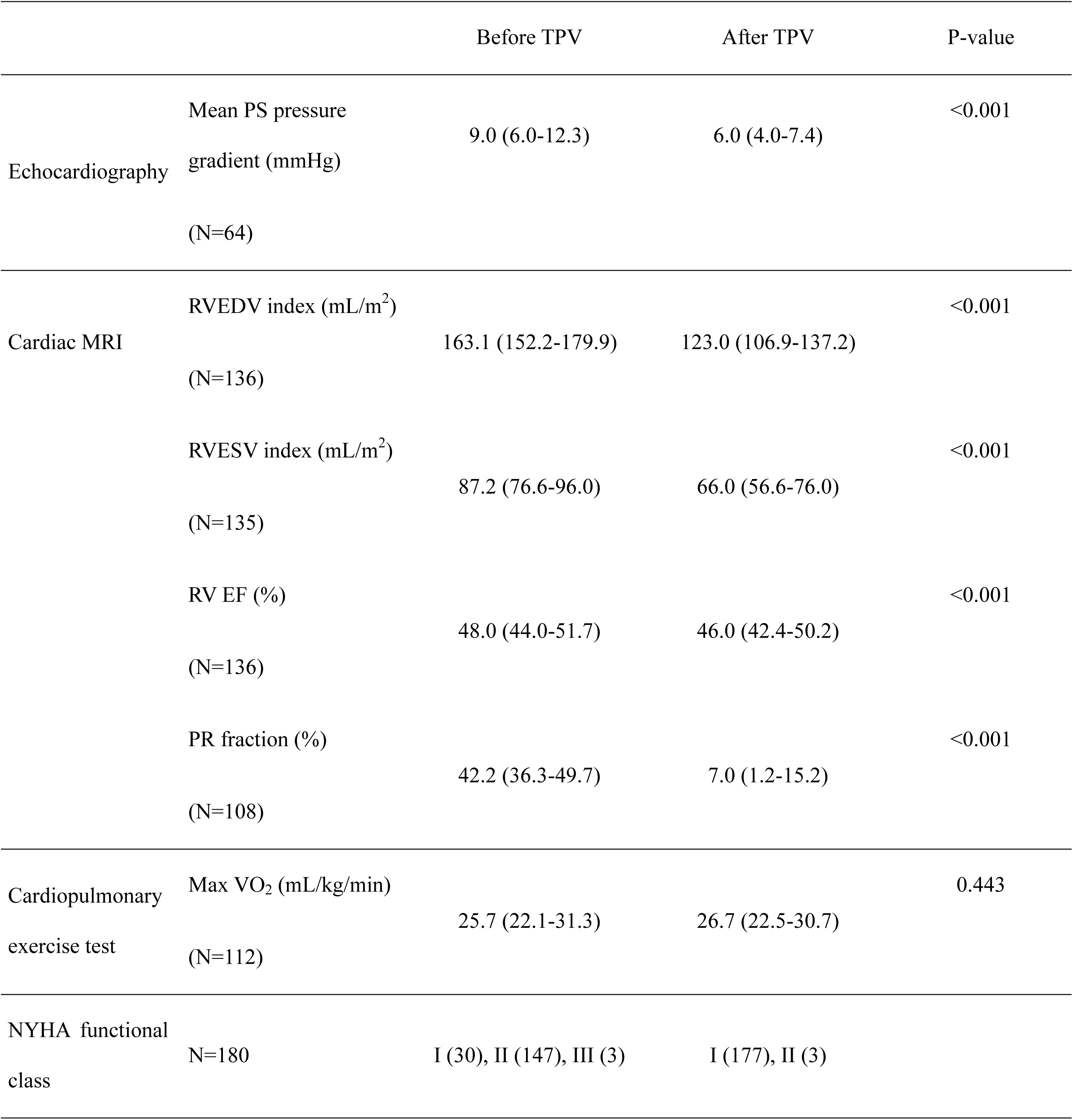
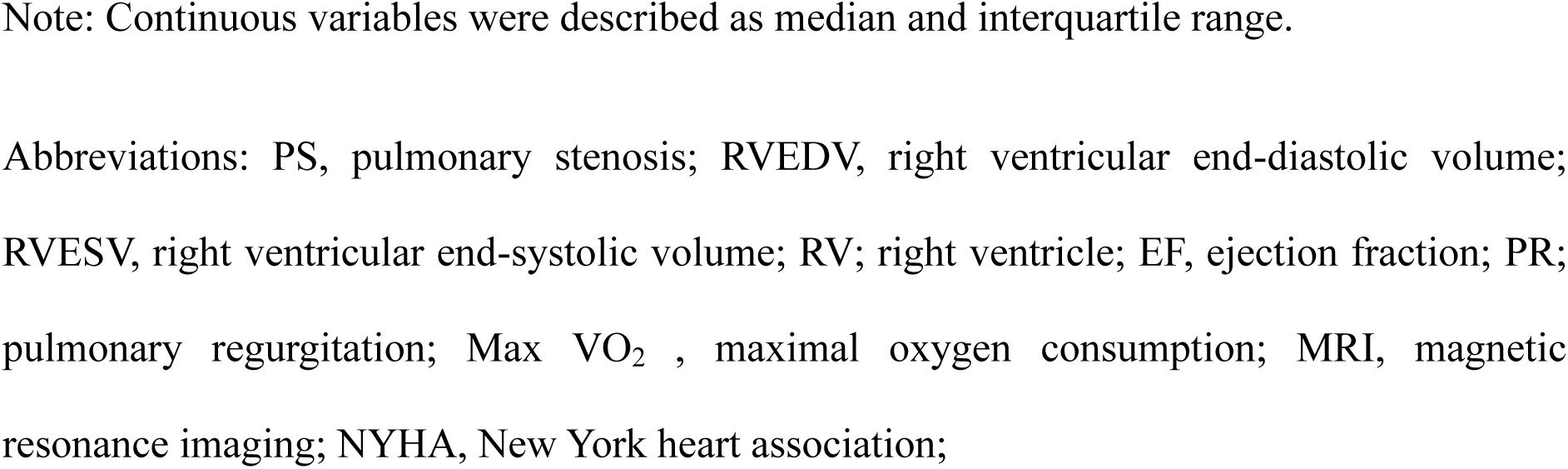
Hemodynamic and Clinical Changes after Pulsta Valve Implantation.

No adverse events, such as infective endocarditis, stent strut fracture, severe vessel access-related events, pulmonary trauma, or tricuspid valve-related problems, occurred during the follow-up period after Pulsta valve implantation. However, in one case with Type 3 main PA morphology, the Pulsta valve was initially implanted at the mid part of the main PA but resulted in severe paravalvar leakage, leading to moderate PR with a measured PR fraction of 33.4%. Consequently, the patient underwent a redo-PPVI one year later, with the Pulsta valve placed at the proximal part of the main PA to resolve the issue.

## Discussion

The introduction of new valves and technical innovations has expanded the indications for PPVI, now encompassing patients with large native RVOT conditions.^7,12,13^ In patients with large native RVOT undergoing PPVI, the insertion site should be individualized based on the anatomical characteristics and the size of the main PA because of diversity of native RVOT. The anatomical features of the main PA has been classified into five types in general (Pyramidal, Straight, Reverse pyramidal, Convex, Concave).^11^ Patients with a pyramidal shape main PA should be implanted at the distal part of the main PA. For the remaining patients, the Pulsta valve should be implanted in the proximal, mid or distal part of the main PA, depending on the specific anatomical features and the size of the main PA.

The Pulsta valve demonstrates several favorable characteristics compared to other self-expandable valves used for PPVI in the pulmonic position. Firstly, the stent frame of the Pulsta valve has uncovered distal ends, allowing for their positioning without impeding branch PA blood flow, even if they extend into the branch pulmonary arteries. This unique design minimizes the risk of flow obstruction and associated complications. Furthermore, the Pulsta valve exhibits a compact structure, with diameters ranging from 18 to 32 mm in 2-mm increments and a total length of 28 to 38 mm, tailored to fit the average length of the main PA which provide low risk of ventricular arrhythmia after valve implantation. Additionally, both ends of the valve are flared by an extra 4mm beyond the outer diameter. These rather compact and short characteristics enable the Pulsta valve to adapt to diverse RVOT geometries while reducing the occurrence of premature ventricular contractions or ventricular tachycardia following the procedure by minimizing RVOT muscle area. Historically, post-procedural arrhythmias, including ventricular tachycardia, have been a concern with PPVI^14^, but the unique structural features of the Pulsta valve have alleviated this concern. Another advantage of the Pulsta valve lies in its cylindrical shape, distinguishing it from other self-expandable valves such as the Harmony, Alterra, Med-Zenith PT, and Venous-P valves, which exhibit an hourglass shape.^15^ In addition, the Pulsta valve utilizes a knitted (woven texture) nitinol wire backbone, a characteristic that contributes to its low risk of stent fracture. Fracture of the stent is a significant complication that can occur following PPVI, with reported rates of up to 4.4%.^16–19^ However, it is noteworthy that during the mid-term follow-up period, no instances of stent fracture were observed with the Pulsta valve. Finally, the Pulsta valve can be loaded directly using a low-profile (18 or 20 Fr) catheter and introduced into access vein directly, thereby minimizing the potential for damage to the accessed venous route and manual compression at the puncture site is sufficient for hemostasis. The above-mentioned features make the Pulsta valve user-friendly for performing PPVI because the success of the PPVI procedure depends heavily on the appropriate selection of the valve size and accurate placement of the Pulsta valve within the designated landing zone.

The target Pulsta valve insertion site according to each main PA shape are described in Figure 4. The target area for PPVI in the RVOT is typically the main PA. However, native RVOT patients may present with various anatomical substrates that pose challenges to the procedural success. Historically, Type 1 RVOT shape was considered unsuitable for PPVI with balloon-expandable valves due to the high risk of proximal migration of the implanted prosthesis.^11^ To overcome this obstacle, various techniques such as the Russian doll technique, PA jailing technique, off-pump surgical main PA banding and Melody valve implantation, RVOT pre-stenting and large SAPIEN valve implantation have been introduced.^20–24^ Pulsta valve, with its maximal total length of less than 38 mm that fits the average length of the main PA, was specifically developed to address this concern. Its unique structure enables advancement to the distal part of the main PA, thereby expanding the indications for PPVI to include patients with Type 1 RVOT. Additionally, Type 3 morphology, characterized by a reverse pyramidal shape, has been also considered unsuitable for PPVI due to the increased risk of valve instability. Adequate landing zone is crucial in such cases. In this study, all patients with Type 1 and Type 3 morphology underwent PPVI using the 38 mm length Pulsta valve to mitigate the risk of valve instability.

**Figure 4.**
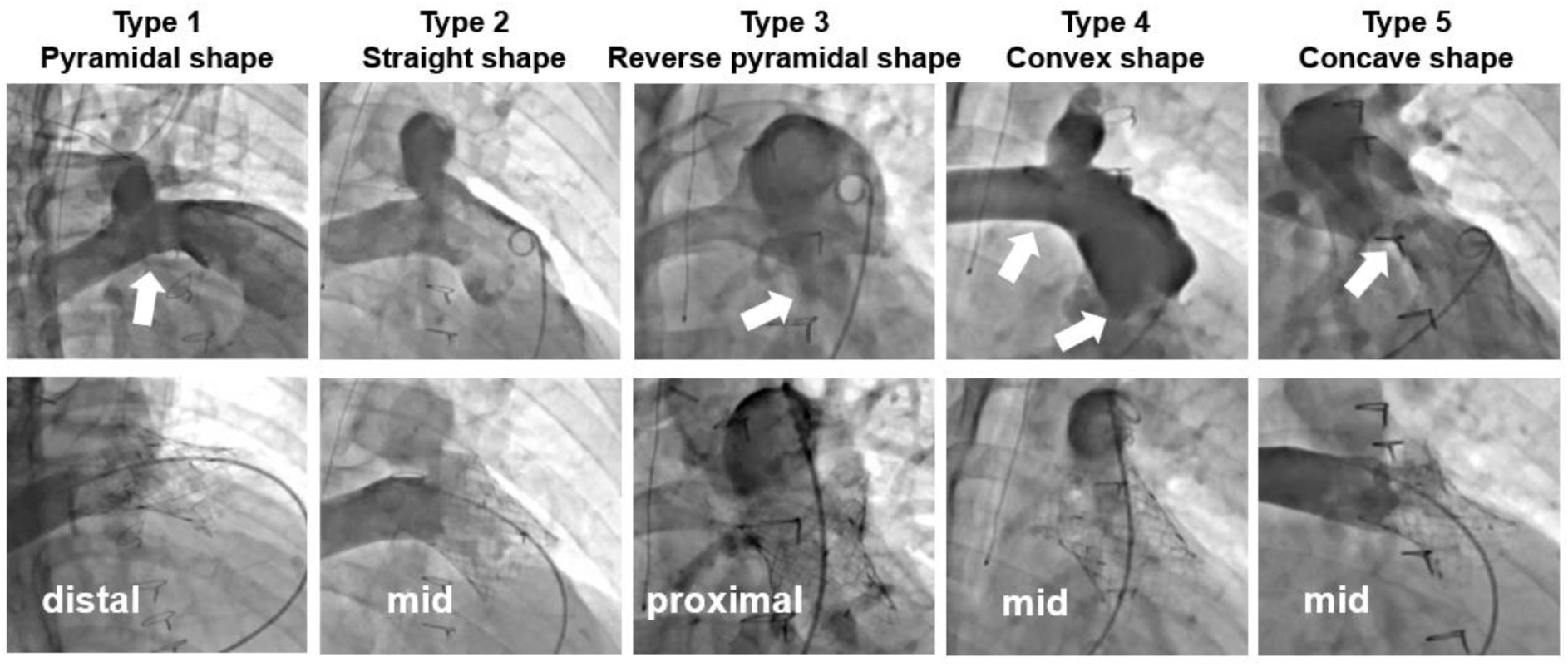
Target Pulsta valve insertion site based on the shape of each main pulmonary artery. The right anterior oblique and cranial view allows for the assessment of the branch pulmonary artery bifurcation, aiding in the selection of the target Pulsta valve insertion site. White arrow indicates the narrowest area for the main Pulsta valve landing zone.

For Types 2 and 4 morphology, the mid portion of the main PA is the preferred landing zone, although placement in the proximal or distal portions of the main PA is also tolerable. In Type 4 morphology, which is associated with a higher risk of paravalvular leakage, if concerns regarding paravalvular leakage arise, slight oversizing of the valve is acceptable.^25^ In Type 3 morphology, the Pulsta valve should be implanted in the proximal or mid part of the main PA, depending on the specific anatomical features and size of the main PA. Type 5 morphology indicates that the mid portion of the main PA is the most suitable landing zone for PPVI.

The Pulsta valve has several mentioned advantages, but it also has limitations. Currently, the Pulsta valve is developed only up to a diameter of 32 mm, making the procedure feasible for patients with a main PA diameter up to 30 mm. For patients with an expanded diameter beyond 30 mm, a larger-sized valve is necessary.

This study has several limitations. Firstly, being a retrospective study, it is inherently constrained by the limitations associated with this design. Secondly, as a multi-center clinical study, it is limited by the relatively small number of patients and follow-up duration is still rather short. However, it’s worth noting that despite these limitations, this study stands as the largest among reported studies examining the outcomes of the Pulsta valve up to the present.

In conclusion, Pulsta valve showed adaptability to the various morphological characteristics of each patient’s main PA by tailored insertion site selection during PPVI by its distinct advantages of compact and user-friendly design. These features have expanded the indications for PPVI to include diverse native RVOT patients. By considering the unique anatomical features of main PA and using the Pulsta valve, the success rate of PPVI can be improved, leading to better outcomes for native RVOT patients requiring PPVI.

## Data Availability

The datasets used and analysed during the current study available from the corresponding author on reasonable request.

## Acknowledgements

This research was supported by SNUH Lee Kun-hee Child Cancer & Rare Disease Project, Republic of Korea (grant number: 23C0220100).

## Source of Funding

This research was funded by SNUH Lee Kun-hee Child Cancer & Rare Disease Project, Republic of Korea (grant number: 23C0220100).

## Disclosures

The authors have no conflicts of interest to disclose.

## Supplemental materials Supplement video 1

The Pulsta valve leaflet movement during systole and diastole. The Pulsta valve is a tri-leaflet valve using treated porcine pericardium which is basically open state.

## Notes

### Competing Interest Statement

The authors have declared no competing interest.

### Author Declarations

This study was approved by the Institutional Review Board of the Seoul National University Hospital (IRB number: 2209-130-1362).

